# Epidemiological differences in the impact of COVID-19 vaccination in the United States and China

**DOI:** 10.1101/2021.01.07.21249380

**Authors:** Monia Makhoul, Hiam Chemaitelly, Houssein H. Ayoub, Shaheen Seedat, Laith J. Abu-Raddad

**Affiliations:** Infectious Disease Epidemiology Group, Weill Cornell Medicine-Qatar, Cornell University, Qatar Foundation - Education City, Doha, Qatar; World Health Organization Collaborating Centre for Disease Epidemiology Analytics on HIV/AIDS, Sexually Transmitted Infections, and Viral Hepatitis, Weill Cornell Medicine–Qatar, Cornell University, Qatar Foundation – Education City, Doha, Qatar; Department of Population Health Sciences, Weill Cornell Medicine, Cornell University, New York City, New York, USA; Department of Mathematics, Statistics, and Physics, Qatar University, Doha, Qatar

**Keywords:** SARS-CoV-2, COVID-19, coronavirus, epidemiology, vaccine, mathematical model

## Abstract

**Background:** The objective of this study was to forecast the impact of COVID-19 vaccination in the United States (US) and China, two countries at different epidemic phases.

**Methods:** A mathematical model describing SARS-CoV-2 transmission and disease progression was used to investigate vaccine impact. Impact was assessed both for a vaccine that prevents infection (*VE*_*S*_ = 95%) and a vaccine that prevents only disease (*VE*_*P*_ = 95%).

**Results:** For *VE*_*S*_ = 95% and gradual easing of restrictions, vaccination in the US reduced the peak incidence of infection, disease, and death by >55% and cumulative incidence by >32%, and in China by >77% and >65%, respectively. Nearly three vaccinations were needed to avert one infection in the US, but only one was needed in China. For *VE*_*P*_ = 95%, benefits of vaccination were half those for *VE*_*S*_ = 95%. In both countries, the impact of vaccination was substantially enhanced with rapid scale-up, vaccine coverage >50%, and slower or no easing of restrictions, particularly in the US.

**Conclusions:** COVID-19 vaccination can flatten, delay, and/or prevent future epidemic waves. However, vaccine impact is destined to be heterogeneous across countries because of an underlying “epidemiologic inequity” that reduces benefits for countries already at high incidence, such as the US. Despite 95% efficacy, actual vaccine impact could be meager in such countries, if vaccine scale-up is slow, acceptance of the vaccine is poor, or restrictions are eased prematurely.

**One Sentence Summary:** Vaccine impact will be heterogeneous across countries disadvantaging countries at high incidence. This heterogeneity can be alleviated with rapid vaccination scale-up and limited easing of restrictions.

## Introduction

With over 80 million infections and a death toll approaching two million [1], the severe acute respiratory syndrome coronavirus 2 (SARS-CoV-2) pandemic has been one of the most challenging global health emergencies in recent history [2]. The unparalleled burden on healthcare systems has necessitated unprecedented restrictions on mobility and on social and economic activities [3,4]. The ensuing losses to national and global economies are probably the largest since the Great Depression [2,5].

We previously developed a mathematical model to investigate the generic population-level impact of SARS-CoV-2 vaccination [6]. In light of recently produced vaccines with ∼95% efficacy against Coronavirus Disease 2019 (COVID-19) symptomatic disease [7,8], the model was extended to assess the impact of these novel vaccines on COVID-19 morbidity and mortality in two major countries at different epidemic phases, the United States (US) and China. The impact was assessed under two different assumptions for the mechanism of action of the vaccine, that it prevents both infection and disease, or that it prevents only disease. The impact was further assessed at different levels of vaccine coverage, different time courses for vaccine scale-up, and different schedules for easing of social and physical distancing restrictions, following the launch of vaccination.

## Materials and Methods

### Mathematical model and parameterization

The extended model was age-structured, stratifying the population into cohorts based on vaccination status, age group, infection status, infection stage, and disease stage. Population movement among cohorts was determined using a set of coupled nonlinear differential equations. Given interest in assessing vaccination impact in the short-term (over only 2021), demography was assumed stable. Contact between individuals in different age groups was determined using an age-mixing matrix that allowed a range of assortativeness in mixing. Details of the model are in Supplementary Information Texts S1A-S1B and Figures S1-S2. The model was coded, fitted, and analyzed using MATLAB R2019a [9].

Since the evidence suggests that reinfection with this virus is a rare event [10-14], those recovered from infection were assumed protected against reinfection, but only for one year, based on the behavior of other “common cold” coronaviruses [15]. For the same purpose, it was assumed that vaccine-induced immunity will also last for only one year. The waning of both natural and vaccine immunity was assumed to follow a gamma distribution of order *n* = 15. That is, most people lose their immunity after about one year, and only a small minority lose their immunity after a period that is either much shorter or much longer than one year (Figure S3).

The model was parameterized using state-of-the-art empirical evidence for the infection’s natural history and epidemiology. The distribution of infected individuals across the mild (or asymptomatic), severe, or critical infection stages and the infection mortality rate in each age group were based on the analyzed epidemic of France [16]. All age groups were assumed (biologically) equally susceptible to this infection. Population demographic information (size, age distribution, and life expectancy) were obtained from the United Nations World Population Prospects database [17]. Details of model parameters, values, and justifications are in Supplementary Information Text S1C and Tables S1-S2.

### Characteristics of the vaccine and scale-up scenarios

Since the primary endpoint of the vaccine’s randomized clinical trials was efficacy of the vaccine against laboratory-confirmed COVID-19 cases [7,8,18], and not just *any* infection, documented or undocumented, it is unknown whether the vaccine prophylactically reduces susceptibility to the infection (that is, *VE*_*S*_ efficacy defined as the proportional reduction in the susceptibility to infection among those vaccinated, compared to those unvaccinated [6]), or whether it just reduced serious symptomatic COVID-19 cases with no effect on infection (that is, *VE*_*P*_ efficacy against disease progression, defined as the proportional reduction in the fraction of individuals with severe or critical infection among those vaccinated, but who still acquired the infection, compared to those unvaccinated [6]). These two mechanisms of action bracket the two extremes for the vaccine’s biological effect, with the former mechanism being the most optimistic (reducing both infection and disease) and the latter being the most pessimistic (reducing only disease).

Notwithstanding this uncertainty, the impact of the vaccine was assessed under each of these mechanisms of action, assuming *VE*_*S*_ = 95% or *VE*_*P*_ = 95%. In the baseline scenario, the vaccine was introduced in both countries on January 1, 2021 with a scale-up to reach vaccine coverage of 80% by the end of 2021. Given that the purpose of vaccination is to alleviate the need for restrictions that have stifled social and economic activities, social distancing restrictions were assumed to be eased gradually over six months, so that “normalcy” would be attained at the end of these six months. Normalcy was defined as a social contact rate in the population equal to that prior to the pandemic.

Since the US has experienced a large epidemic, it was assumed that 20% of the US population has already been infected by January 1, 2021, with those already infected (if subsequently vaccinated) not benefiting from the immunity conferred by this vaccine. Moreover, the basic reproduction number at time of onset of vaccination was assumed at *R*_0_ = 1.2, reflecting the current phase of an expanding epidemic. It was also assumed that *R*_0_ will gradually increase with easing of restrictions to reach *R*_0_ = 4 at the end of six months. The value of *R*_0_ = 4 is justified by existing estimates assuming a “natural” epidemic in the absence of interventions [19,20].

For China, it was assumed that the entire population is still susceptible to the infection, given the small number of documented infections relative to its large population size, and that the epidemic was contained [1,21]. Moreover, it was assumed that *R*_0_ = 1 at the onset of vaccination (reflecting the non-expanding epidemic), but that *R*_0_ will gradually increase with easing of restrictions to reach an *R*_0_ = 4 at the end of six months.

### Measures of vaccine impact

The population-level impact of SARS-CoV-2 vaccination was assessed by quantifying incidence, cumulative incidence, and reduction in incidence of infections, severe disease cases, critical disease cases, and COVID-19 deaths arising in the presence of vaccination compared to the counter-factual scenario of no-vaccination. Vaccine *effectiveness*, that is number of vaccinations needed to avert one infection or one adverse disease outcome (ratio of the number of vaccinations relative to the number of averted outcomes) was further calculated to inform future cost-effectiveness analyses.

### Uncertainty analysis

A multivariable uncertainty analysis was conducted to determine the range of uncertainty for model predictions using five-hundred model runs. At each run, Latin Hypercube sampling [22,23] was applied to select vaccine efficacy from within its reported credible range [8], and to select a vaccine duration of protection within ±30% of one-year duration. The resulting distribution for vaccine impact across all 500 runs was used to calculate the predicted means of different outcomes and the uncertainty associated with those means.

## Results

For *VE*_*S*_ = 95%, vaccination in the US flattened the epidemic curve, but did not prevent a new epidemic wave, though it resulted in a smaller one, with the assumed gradual easing of restrictions following the onset of vaccination (Figure 1). The vaccine reduced peak incidence of infection, severe disease, critical disease, and COVID-19 death by 59.6%, 59.5%, 59.0%, and 55.3%, respectively, and the cumulative number of infections, severe disease cases, critical disease cases, and deaths by 35.7%, 35.2%, 35.0%, and 32.7%, respectively, by end of 2021. However, incidence started to increase toward the end of 2021, as vaccine immunity waned and those previously infected began losing their protective immunity against reinfection.

**Figure 1.**
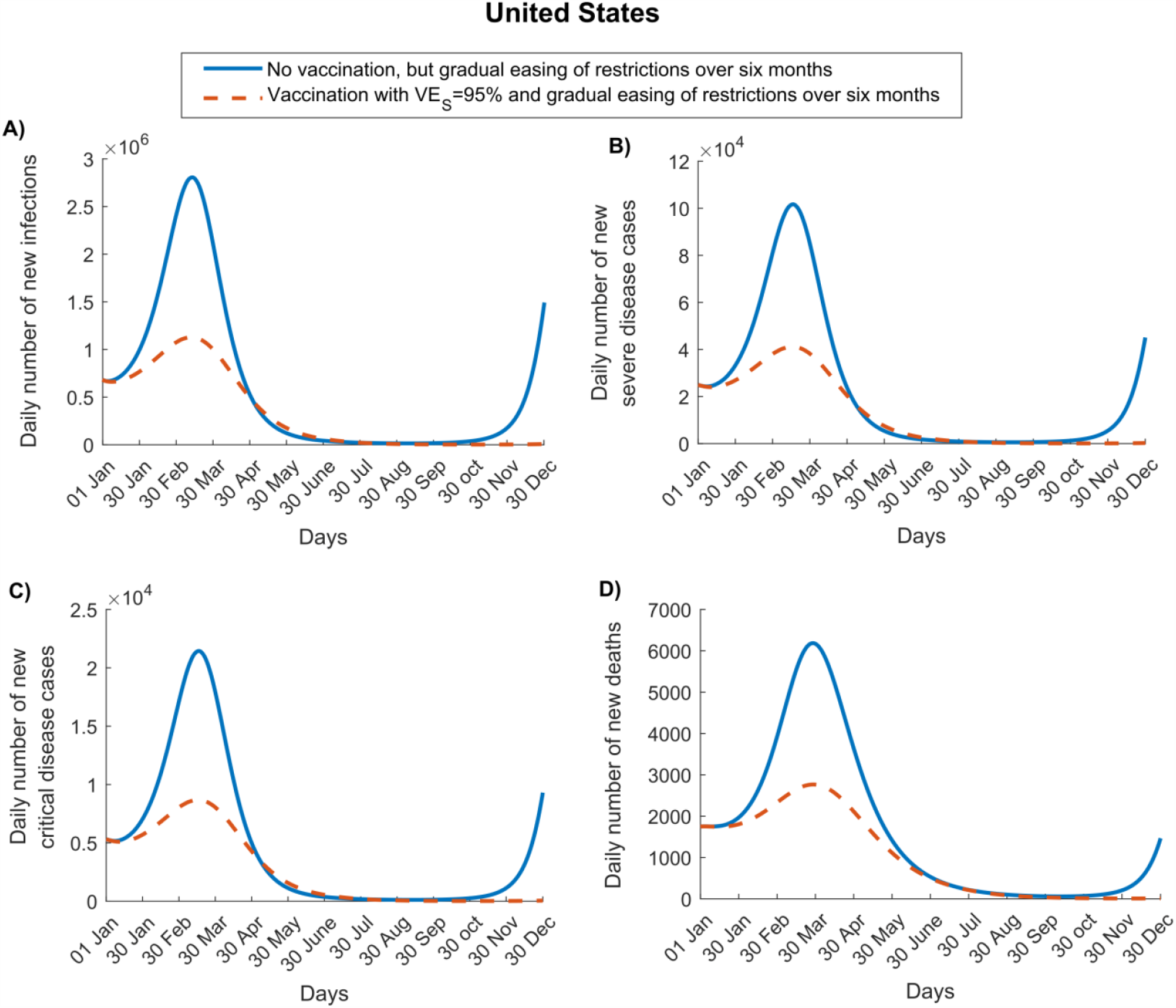
Impact of SARS-CoV-2 vaccination on numbers of A) new infections, B) new severe disease cases, C) new critical disease cases, and D) new deaths in the United States. The vaccine is assumed to have an efficacy of 95% against infection and is introduced on January 1, 2021, when the proportion of the population already infected is 20%. Vaccine coverage is scaled up to reach 80% by December 31, 2021. Duration of both vaccine protection and natural immunity is one year. This scenario assumes an *R*_0_ of 1.2 on January 1, 2021, which increases with gradual easing of restrictions to reach 4.0 after six months. Results of a scenario assuming that the vaccine has no efficacy against infection, but an efficacy of 95% against severe and critical disease are shown in Figure S4.

For *VE*_*P*_ = 95%, the vaccination had no impact on infection (as it does not protect against infection) and less impact on disease and death (Figure S4). Peak incidence of severe disease, critical disease, and death was reduced by only 22.0%, 22.0%, and 21.1%, respectively. The cumulative number of severe disease cases, critical disease cases, and deaths were reduced by only 17.4%, 17.2%, and 16.7%, respectively, by end of 2021.

In China, the impact of vaccination was larger than in the US, as the vaccine was introduced at a time when disease incidence was negligible. For *VE*_*S*_ = 95%, vaccination not only flattened the epidemic curve, but also delayed it by a few months (Figure 2). The vaccine reduced peak incidence of infection, severe disease, critical disease, and death by 85.6%, 84.2%, 84.3%, and 77.3%, respectively, and the cumulative number of infections, severe disease cases, critical disease cases, and deaths by 65.7%, 65.0%, 65.3%, and 65.3%, respectively, by end of 2021.

**Figure 2.**
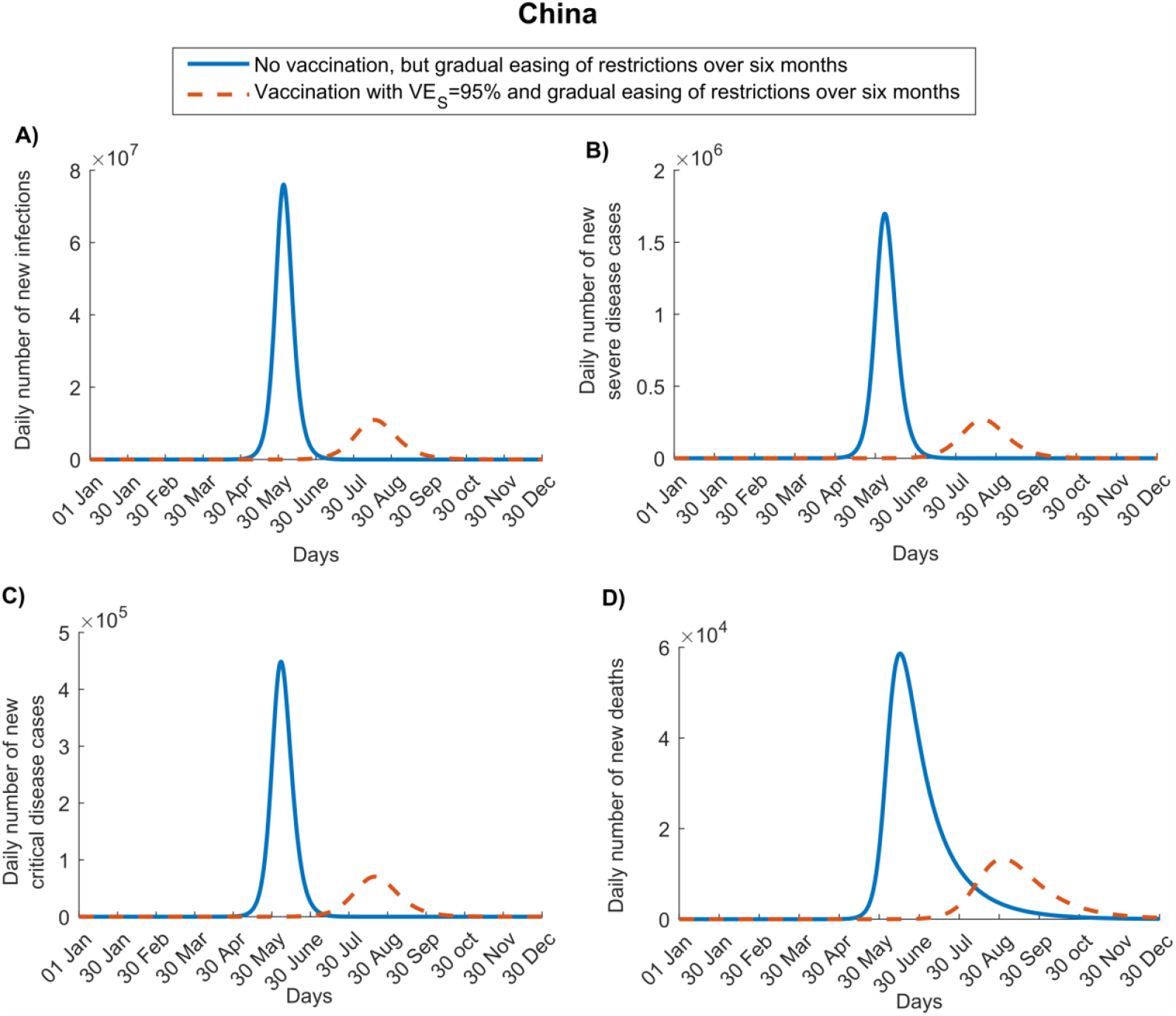
Impact of SARS-CoV-2 vaccination on numbers of A) new infections, B) new severe disease cases, C) new critical disease cases, and D) new deaths in China. The vaccine is assumed to have an efficacy of 95% against infection and is introduced on January 1, 2021. Vaccine coverage is scaled up to reach 80% by December 31, 2021. Duration of both vaccine protection and natural immunity is one year. This scenario assumes an *R*_0_ of 1.0 on January 1, 2021, which increases with gradual easing of restrictions, to reach 4.0 after six months. Results of a scenario assuming that the vaccine has no efficacy against infection, but an efficacy of 95% against severe and critical disease are shown in Figure S5.

For *VE*_*P*_ = 95%, the vaccination had less impact on disease and death (Figure S5). Peak incidence of severe disease, critical disease, and death was reduced by 44.5% for all of these indicators, as well as for their cumulative numbers.

In the US, for *VE*_*S*_ = 95%, the cumulative number of averted disease cases increased steadily in response to shorter scale-up (to 80% coverage) (Figure 3A). However, in China, there was no additional benefit to be had by shortening scale-up to less than 8 months, as the epidemic was fully contained (Figure 3C). Similar results were obtained for *VE*_*P*_ = 95%, as shown for the US and China (Figures S6A and S6C), respectively.

For *VE*_*S*_ = 95%, the cumulative number of averted disease cases increased steadily with higher vaccine coverage (by end of 2021) in both countries (Figures 3B and 3D). The gains in averted disease cases increased sharply as vaccine coverage exceeded 70% in the US and 50% in China, because such coverage prevented a much larger epidemic wave. Similar results were obtained for *VE*_*P*_ = 95%, in both the US and China (Figures S6B and S6D, respectively).

**Figure 3.**
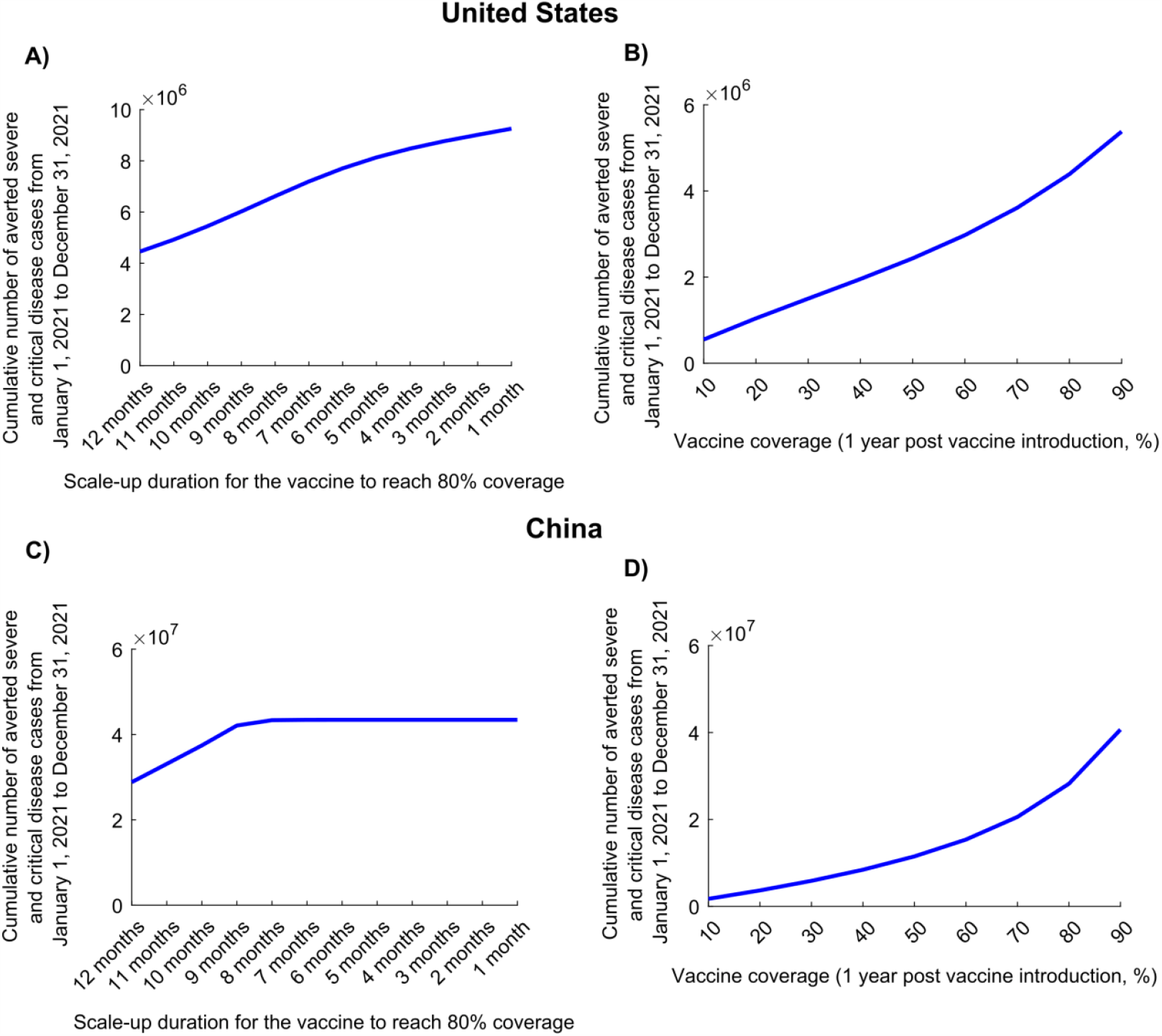
Impact of vaccine scale-up duration and vaccine coverage on numbers of averted severe and critical disease cases for a vaccine that protects against both infection and disease. Cumulative averted severe and critical disease cases in A) the United States and C) China at different vaccination scale-up durations to reach 80% coverage. Cumulative averted severe and critical disease cases in B) the United States and D) China at varying levels of vaccine coverage. The vaccine is assumed to have an efficacy of 95% against infection and is introduced on January 1, 2021, when the cumulative proportion of the population infected is 20% in the United States and 0% in China. Duration of both vaccine protection and natural immunity is one year. This scenario assumes gradual easing of restrictions within 6 months. The results of a scenario assuming the vaccine has no efficacy against infection, but an efficacy of 95% against severe and critical disease is shown in Figure S6.

In the US, for *VE*_*S*_ = 95%, the effectiveness of the vaccine in preventing infection (Figure 4A), severe disease (Figure 4B), critical disease (Figure 4C), and death (Figure 4D), was substantially enhanced by more rapid scale-up to reach 80% coverage, since the epidemic was already at high incidence at time vaccination was launched. Whereas in the US, only one vaccination was needed to avert one infection, provided that scale-up could be accomplished in 6 months, nearly 3 vaccinations were needed to avert one infection if the scale-up required 12 months. This, however, was not the case in China (Figure S7). Regardless of the speed of scale-up, only one vaccination was needed to avert one infection.

**Figure 4.**
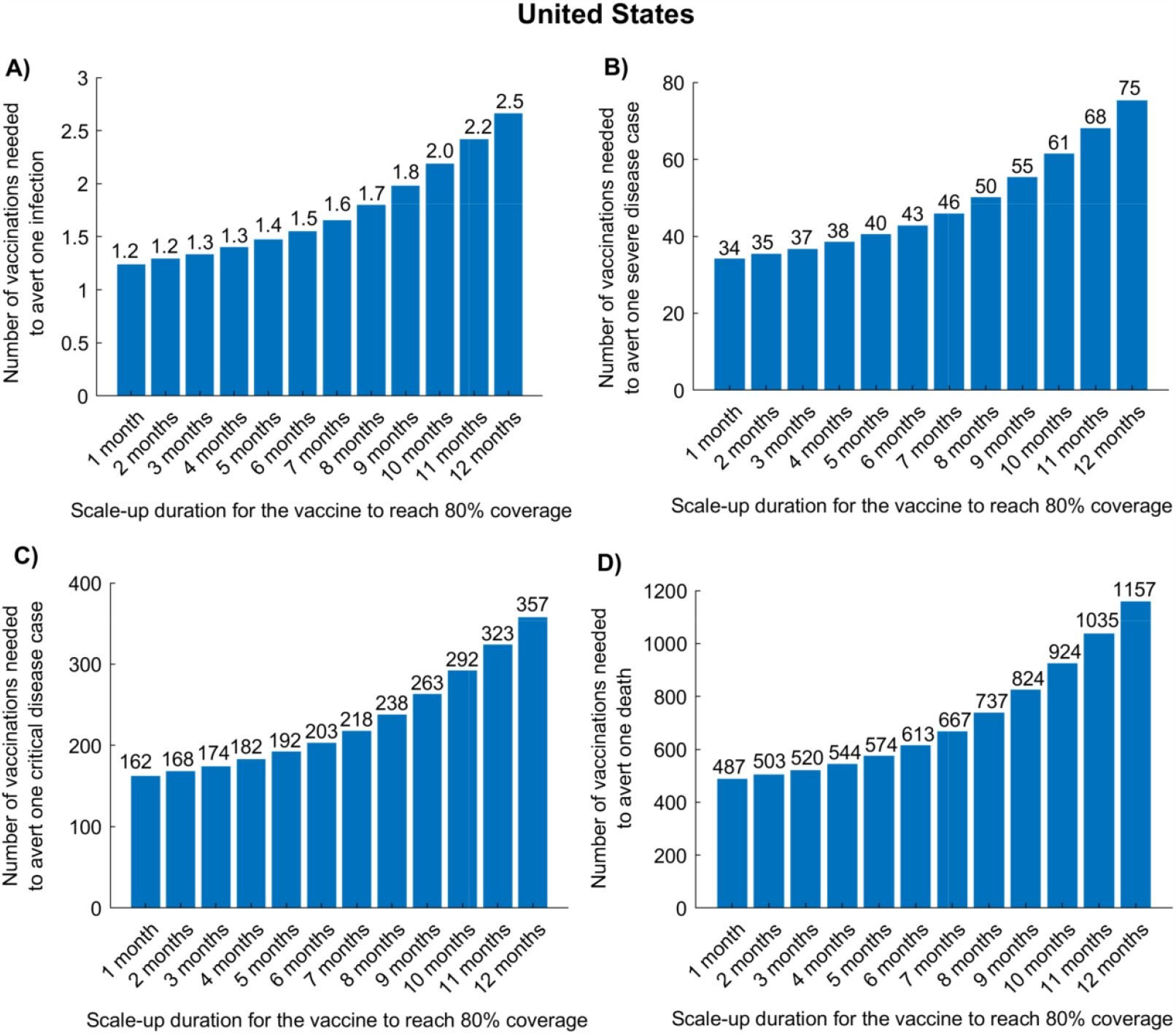
Impact of vaccine scale-up duration on the number of vaccinations needed to avert one infection (A), one severe disease case (B), one critical disease case (C), and one death (D) in the United States. The vaccine is assumed to have an efficacy of 95% against infection and is introduced on January 1, 2021, when the cumulative proportion of the population infected is 20%. Duration of both vaccine protection and natural immunity is one year. This scenario assumes a gradual easing of restrictions within 6 months. Corresponding results for China are shown in Figure S7.

Figure 5 shows a comparison of the impact of vaccine scale-up duration on the number of vaccinations needed to avert one severe disease case (Figure 5A), one critical disease case (Figure 5B), and one death (Figure 5C), in the US, between the assumption of *VE*_*S*_ = 95% and that of *VE*_*P*_ = 95%. As expected, a vaccine that prevents infection (and consequently disease) was superior to a vaccine preventing only disease. That superiority was even greater if scale-up is longer, where twice as many vaccinations were needed to avert each of these outcomes. Similar results were obtained for China (Figure S8).

**Figure 5.**
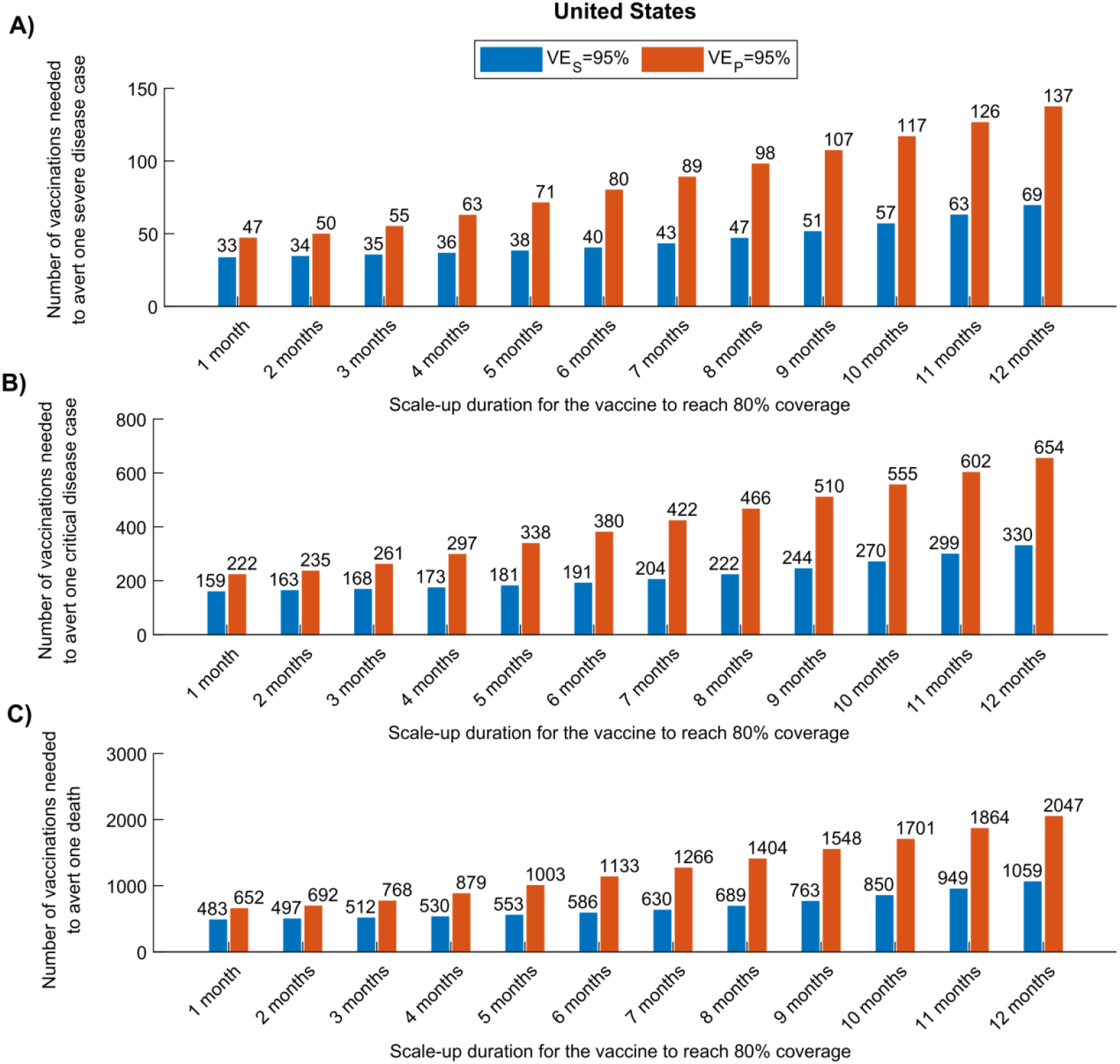
Comparison of the impact of a vaccine acting against infection (*VE*_*S*_ efficacy) versus a vaccine acting only against disease (*VE*_*P*_ efficacy) in the United States. The number of vaccinations needed to avert one severe disease case (A), one critical disease case (B), and one death (C), for a vaccine with *VE*_*S*_ = 95% versus a vaccine with *VE*_*P*_ = 95%. The vaccine is introduced on January 1, 2021, when the cumulative proportion of the population infected is 20%. Duration of both vaccine protection and natural immunity is one year. This scenario assumes a gradual easing of restrictions within 6 months. Corresponding results for China are shown in Figure S8.

In all of the above scenarios, it was assumed that easing of social restrictions would occur during six months following initiation of vaccination. However, as expected, a longer duration for easing restrictions resulted in a more favorable impact of vaccination, in both the US and China (Figure S9).

Uncertainty regarding the projected impact was small in the short-term for the US and China, in the first wave after vaccinations commenced, but it was large toward the end of 2021, as expected, due to uncertainty about persistence of the vaccine’s protective immunity (Figure S10).

## Discussion

The key conceptual finding that emerges from this study is that vaccine impact is strongly dependent on the difference between two essential metrics, “*time to infection*” and “*time to vaccination*.” The competing “hazard” dynamics between the event of infection and the event of vaccination explain the variability of impact under the parameters considered: incidence at the onset of vaccination, duration of scale-up, vaccine coverage, or timing of the easing of restrictions. As the average time to vaccination is shortened *relative to* the average time to infection, by altering these parameters (by more rapid scale-up, slower easing of restrictions, or reducing infection incidence through lockdowns or other restrictive measures), the impact would be more favorable, and fewer vaccinations would be needed to avert one infection or disease outcome.

A striking demonstration of this concept’s importance can be seen in the case of a vaccine that does not prevent infection, but prevents disease with *VE*_*P*_ = 95% (Figures S4 and S5). This vaccine would not affect the time to infection in the population, and any easing of restrictions with vaccination will shorten the time to infection. Accordingly, such a vaccine, despite its 95% efficacy, would end up averting <20% of disease cases and deaths in the US and <50% in China. Since the time to infection in the US is much shorter than in China, as a consequence of the current high incidence rate, vaccine impact will be more favorable in China, where vaccination can be scaled up over a longer duration and still have superior impact to that in the US.

A consequence of the above findings is that vaccine impact will likely be heterogenous among nations. Countries with low or negligible incidence will benefit most from vaccination. Vaccine cost-effectiveness will be also optimized in such countries, with only one vaccination needed to avert one infection for a vaccine with *VE*_*S*_ = 95% (Figure S7).

Several other findings emerged from this study. Vaccination will flatten the epidemic curve, but may not prevent (or delay) a new wave, unless it is scaled up very rapidly (Figures 1-3). There is every virtue in *rapidly* scaling up vaccination, particularly in countries already suffering substantial incidence (Figure 3). Importantly, vaccination impact does not increase linearly with vaccine coverage—gains from vaccination would be proportionally higher if vaccine coverage exceeds 50% (Figure 3), stressing the importance of reaching high vaccine coverage. Easing of restrictions concurrently with vaccination can undermine many benefits of vaccination, as more people are likely to become infected before they are vaccinated. Easing of restrictions needs to be slow and gradual, tailored to the epidemiologic situation in each country (Figure S9). For instance, with its ongoing high incidence, easing of restrictions is not warranted in the US, while vaccination is scaled up.

With 95% efficacy, COVID-19 vaccination is very cost-effective, as fewer than 3 vaccinations are needed to avert one infection, and this effectiveness can be optimized further with more rapid scale-up (Figure 4 and Figure S7). The impact of vaccination in averting disease or death is two-fold higher for a vaccine that prevents infection, compared to a vaccine that only prevents disease (Figures 1-2 versus Figures S4-S5). This is because preventing infection not only prevents disease directly, but also reduces infection circulation; thus, also indirectly reducing disease. Moreover, twice as many vaccinations are needed to avert one disease or death outcome for a vaccine that prevents only disease, compared to one that prevents infection (Figure 5 and Figure S8).

This study has some limitations. Model estimations are contingent on the validity and generalizability of input data. While we used available evidence for SARS-CoV-2 natural history and epidemiology, our understanding of its epidemiology is still evolving. All age groups were assumed equally susceptible to infection, but evidence suggests some biological differences in susceptibility [24-31]. The exact extent of exposure to the infection in both the US and China is unknown, but plays an important role in vaccine impact. From an epidemiological perspective, we assumed that 20% of the US population and a negligible percentage of the Chinese population have been already infected, but vaccine impact can be quite different if such assumptions prove unrealistic. Vaccinated persons were assumed to be immediately protected, once vaccinated, but in reality, vaccine protection develops over the course of a month following inoculation [7,8]. Two parameters remain unknown, despite being critical to the longer-term impact of vaccination: durations of vaccine protection and natural immunity. If both prove to be relatively brief, the impact of the vaccine will be diminished and it may be necessary to periodically re-immunize, or to develop additional vaccines that protect against other circulating strains of this virus.

## Conclusions

COVID-19 vaccination can have an immense impact on averting infection and/or disease. It can substantially flatten and delay future epidemic waves (if not prevent them altogether), and will be highly cost-effective, given the small number of vaccinations needed to avert one infection or one disease outcome. However, the impact of vaccination is likely to vary among countries, reflecting an underlying “epidemiological inequity”, as the epidemic phase in those countries also varies. Nations that will benefit most from vaccination are those where waiting time before vaccination is much shorter than time to infection, that is countries currently at low incidence, such as China. For countries at high incidence, the impact may prove far less than current expectations, despite the vaccine’s 95% efficacy, if vaccination is scaled up slowly and/or if restrictions are eased prematurely. For countries such as the US, there is every virtue in scaling up vaccination rapidly, reaching high vaccine coverage, and delaying any easing of restrictions until viral incidence reaches low levels.

## Data Availability

All data are available in the manuscript and its supplementary materials. Code programmed in MATLAB can be obtained from the authors.

## Author contributions

MM constructed, coded, and parameterized the mathematical model, conducted the analyses, and co-wrote the first draft of the paper. HC supported model parametrization and analyses and co-wrote the first draft of the paper. LJA conceived and led the design of the study and its implementation, and co-wrote the first draft of the paper. All authors contributed to discussion and interpretation of results and to writing of the manuscript. All authors have read and approved the final manuscript.

## Acknowledgments

The authors are grateful for support provided by the Biomedical Research Program and the Biostatistics, Epidemiology, and Biomathematics Research Core, both at Weill Cornell Medicine-Qatar.

## Supplementary Material

### Text S1. SARS-CoV-2 vaccine mathematical model

#### A. Model structure

We extended a recently-developed age-structured deterministic compartmental model [1-5] to describe the impact of vaccination on the severe acute respiratory syndrome coronavirus 2 (SARS-CoV-2) transmission dynamics and progression of the resulting disease, Coronavirus Disease 2019 (COVID-2019), in a given population. The model stratifies the unvaccinated and vaccinated populations into compartments according to age group (0-9, 10-19, 20-29,…, ≥80 years), infection status (uninfected, infected), infection stage (mild, severe, critical), disease stage (severe, critical), and compartments for the gamma distribution (Γ-distribution) describing the waning of natural and vaccine immunity.

Transmission and disease progression dynamics in the vaccinated and unvaccinated cohorts are described in the model using age-specific sets of nonlinear ordinary differential equations, where each age group *a* (*a* = 1, 2,…9) refers to a 10-year age band (0-9,10-19,..70-79) apart from the last group including all those aged ≥0 years. The model is illustrated in Figure S1.

**Figure S1.**
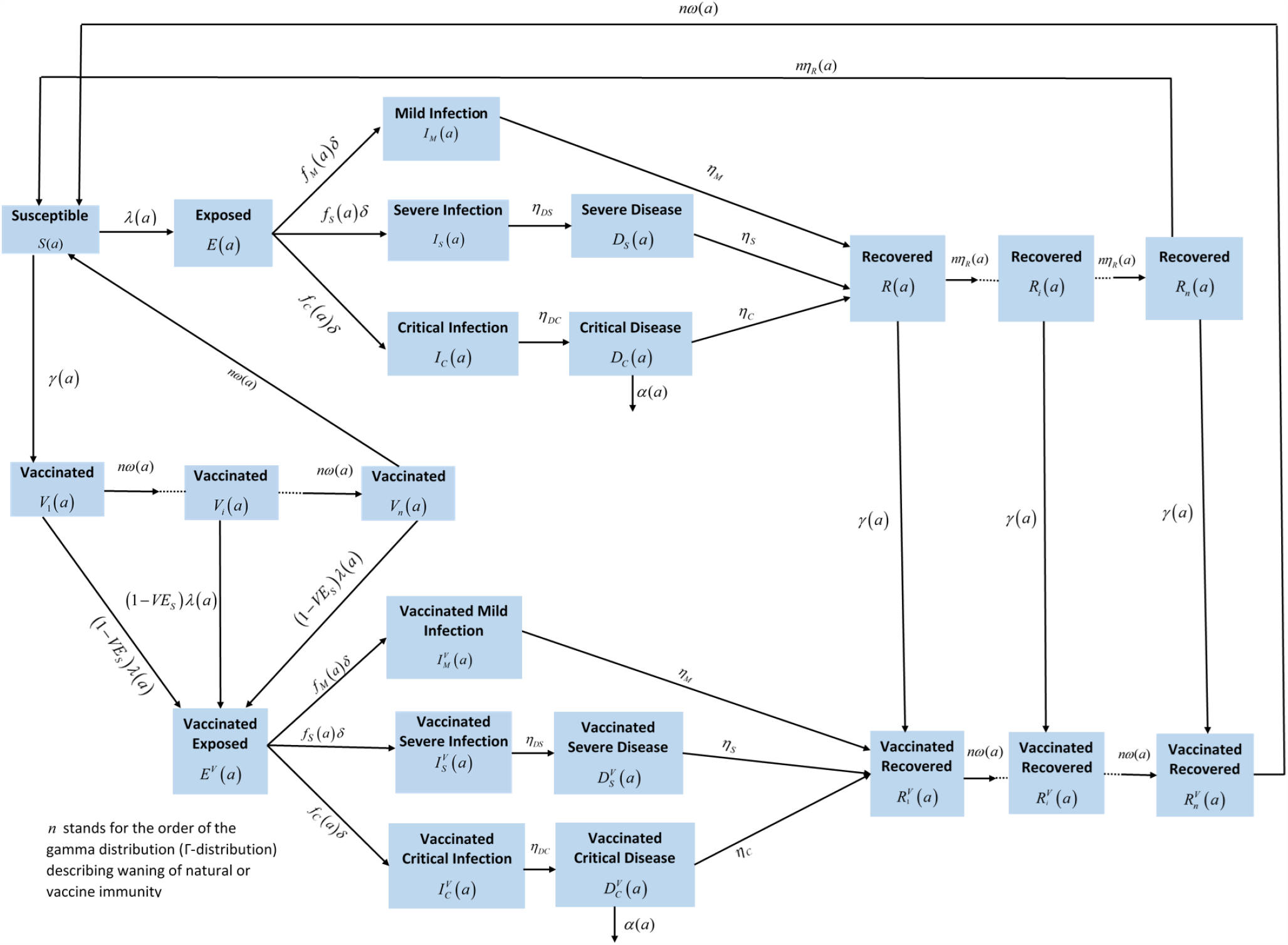
Schematic diagram describing the SARS-CoV-2 transmission dynamics model in presence of a vaccine that reduces susceptibility to infection.

##### Unvaccinated population

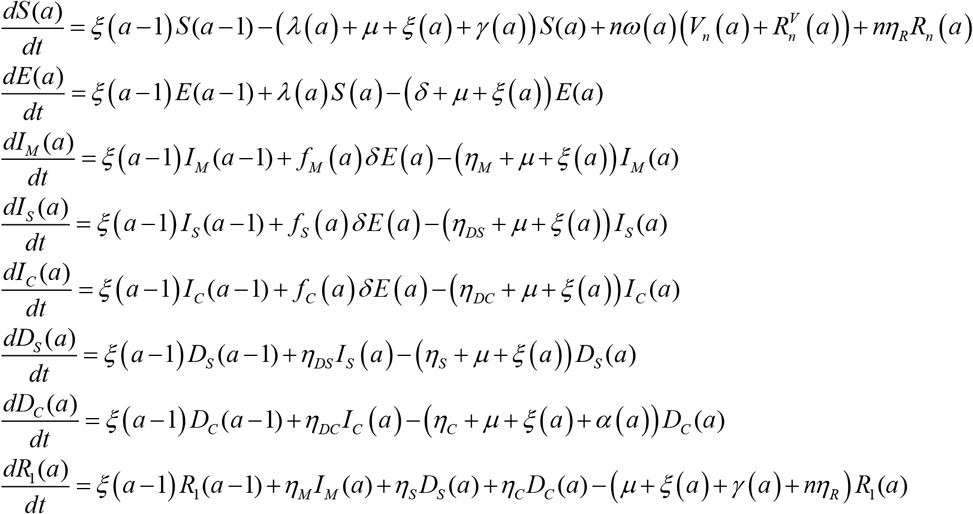

<u>For k=2,..n</u>

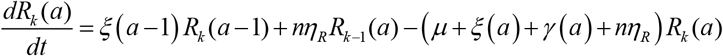

##### Vaccinated populations aged 10+ years

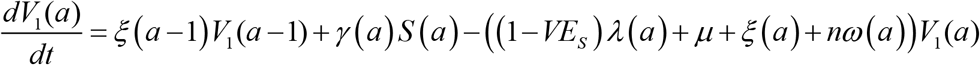

<u>For k=2,..n</u>

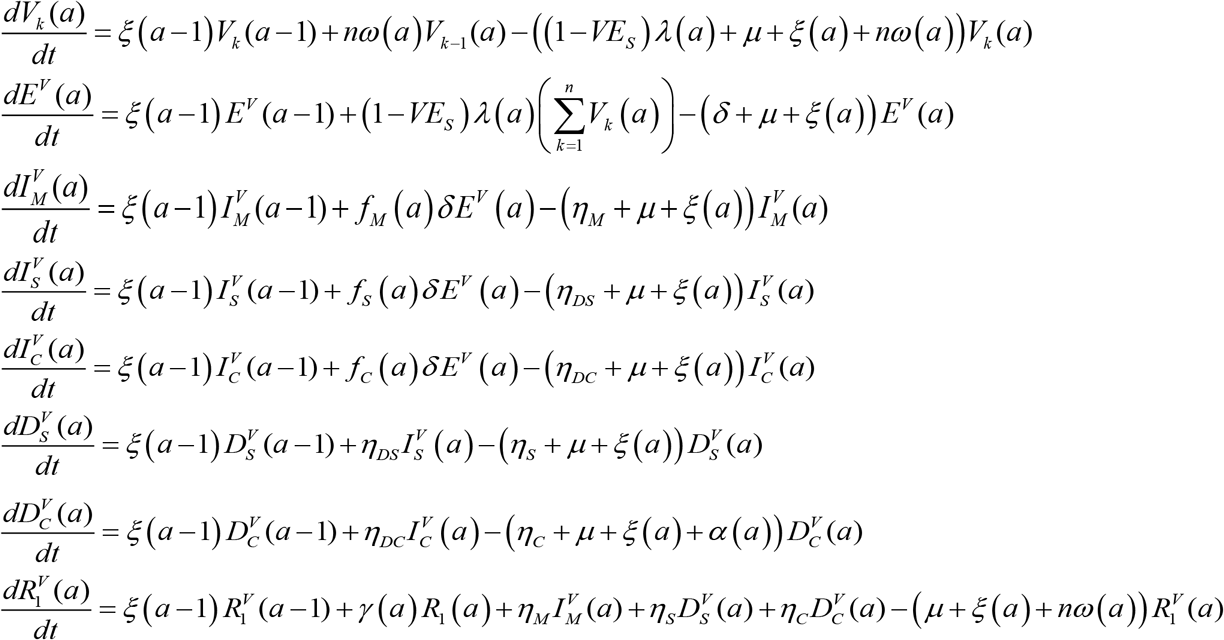

<u>For k=2,..n</u>

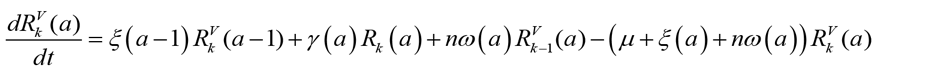

Where *n* stands for the order of the gamma distribution (Γ-distribution) describing waning of natural or vaccine immunity.

The definitions of population variables and symbols used in the equations are in Table S1.

**Table S1.** Definitions of population variables and symbols used in the model.

| Symbol | Definition |
| --- | --- |
| **Transmission dynamics parameters** |  |
| $S(a)$ | Unvaccinated susceptible population |
| $E(a)$ | Unvaccinated latently infected population |
| $I_M(a)$ | Unvaccinated population with asymptomatic or mild infection |
| $I_S(a)$ | Unvaccinated population with severe infection |
| $I_C(a)$ | Unvaccinated population with critical infection |
| $D_S(a)$ | Unvaccinated population with severe disease |
| $D_C(a)$ | Unvaccinated population with critical disease |
| $R_i(a)$ | $i^{th}$ compartment to generate the gamma distribution for the waning of natural immunity among the unvaccinated recovered population |
| $V_i(a)$ | $i^{th}$ compartment to generate the gamma distribution for the waning of vaccine immunity among the vaccinated susceptible population |
| $E^V(a)$ | Vaccinated latently infected population |
| $I_M^V(a)$ | Vaccinated population with asymptomatic or mild infection |
| $I_S^V(a)$ | Vaccinated population with severe infection |
| $I_C^V(a)$ | Vaccinated population with critical infection |
| $D_S^V(a)$ | Vaccinated population with severe disease |
| $D_C^V(a)$ | Vaccinated population with critical disease |
| $R_i^V(a)$ | $i^{th}$ compartment to generate the gamma distribution for the waning of natural immunity among the vaccinated recovered population |
| $N$ | Total population size |
| $n_{age}$ | Number of age groups |
| $n$ | Order of the gamma distribution (Γ-distribution) describing waning of natural or vaccine immunity |
| $\xi(a)$ | Transition rate from one age group to the next age group |
| $\beta$ | Overall infectious contact rate |
| $1/\delta$ | Duration of latent infection |
| $1/\eta_M$ | Duration of asymptomatic or mild infection |
| $1/\eta_{DS}$ | Duration of severe infection infectiousness before isolation and/or hospitalization |
| $1/\eta_S$ | Duration of severe disease following onset of severe disease |
| $1/\eta_{DC}$ | Duration of critical infection infectiousness before isolation and/or hospitalization |
| $1/\eta_C$ | Duration of critical disease following onset of critical disease |
| $1/\eta_r$ | Duration of natural immunity |
| $1/\mu$ | Natural death rate |
| $\alpha(a)$ | Mortality rate in each age group |
| $f_M(a)$ | Proportion of infections that will progress to be mild or asymptomatic infections |
| $f_S(a)$ | Proportion of infections that will progress to be severe infections |
| $f_C(a)$ | Proportion of infections that will progress to be critical infections |
| **Key vaccine product characteristics** |  |
| $VE_s$ | Vaccine efficacy in reducing susceptibility |
| $VE_p$ | Vaccine efficacy in reducing severe and critical disease |
| $1/\omega$ | Duration of vaccine protection |

The force of infection (hazard rate of infection) experienced by the unvaccinated susceptible populations *S* (*a*) is given by

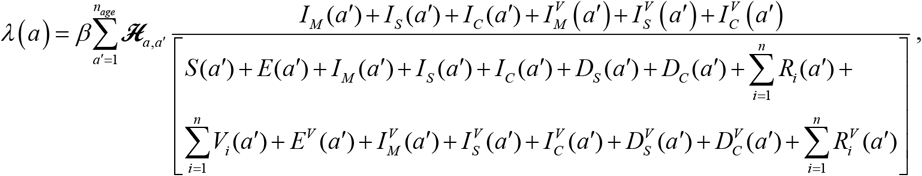

while that experienced by the vaccinated susceptible populations *V* (*a*) is given by

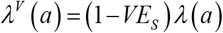

where *β* is the overall infectious contact rate. The mixing among the different age groups is dictated by the mixing matrix ℋ_*a,a*″_. This matrix provides the probability that an individual in the *a* age group will mix with an individual in the *a*′ age group (regardless of vaccination status).

The mixing matrix is given by

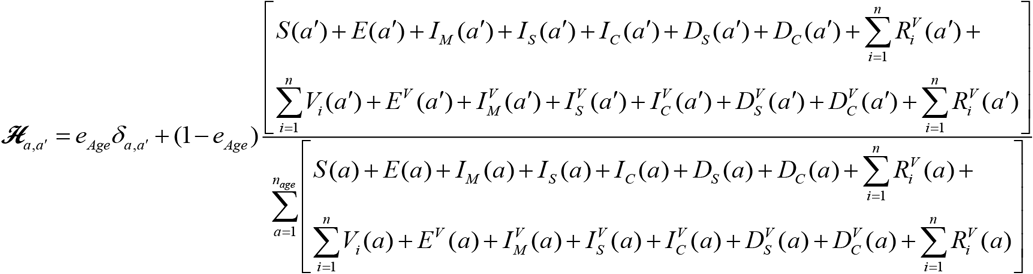

Here, *δ*_*a,a*′_ is the identity matrix. *e*_*Age*_ ∈[0,1] measures the degree of assortativeness in the mixing. At the extreme *e*_*Age*_ = 0, the mixing is fully proportional. Meanwhile, at the other extreme, *e*_*Age*_ = 1, the mixing is fully assortative, that is individuals mix only with members in their own age group.

#### B. Model adjustment for a vaccine that reduces only severe and critical disease

The above model was adjusted to accommodate for a vaccine that has no effect on infection, but that reduces both severe and critical disease with an efficacy *VE*_*P*_. A schematic diagram of the adjusted model is provided in Figure S2.

**Figure S2.**
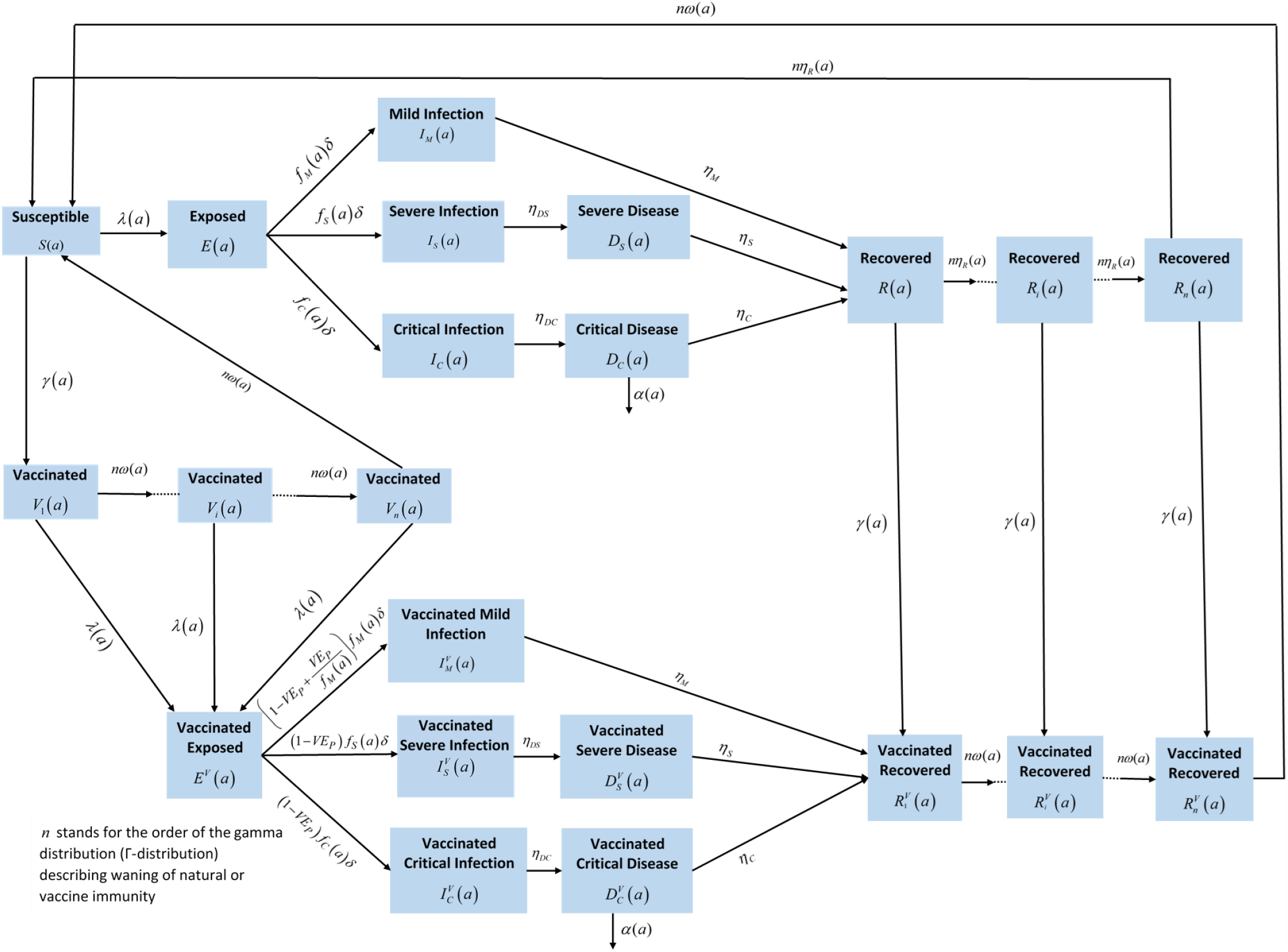
Schematic diagram describing the SARS-CoV-2 transmission dynamics model in presence of a vaccine that reduces severe and critical disease, but does not prevent infection.

**Figure S3.**
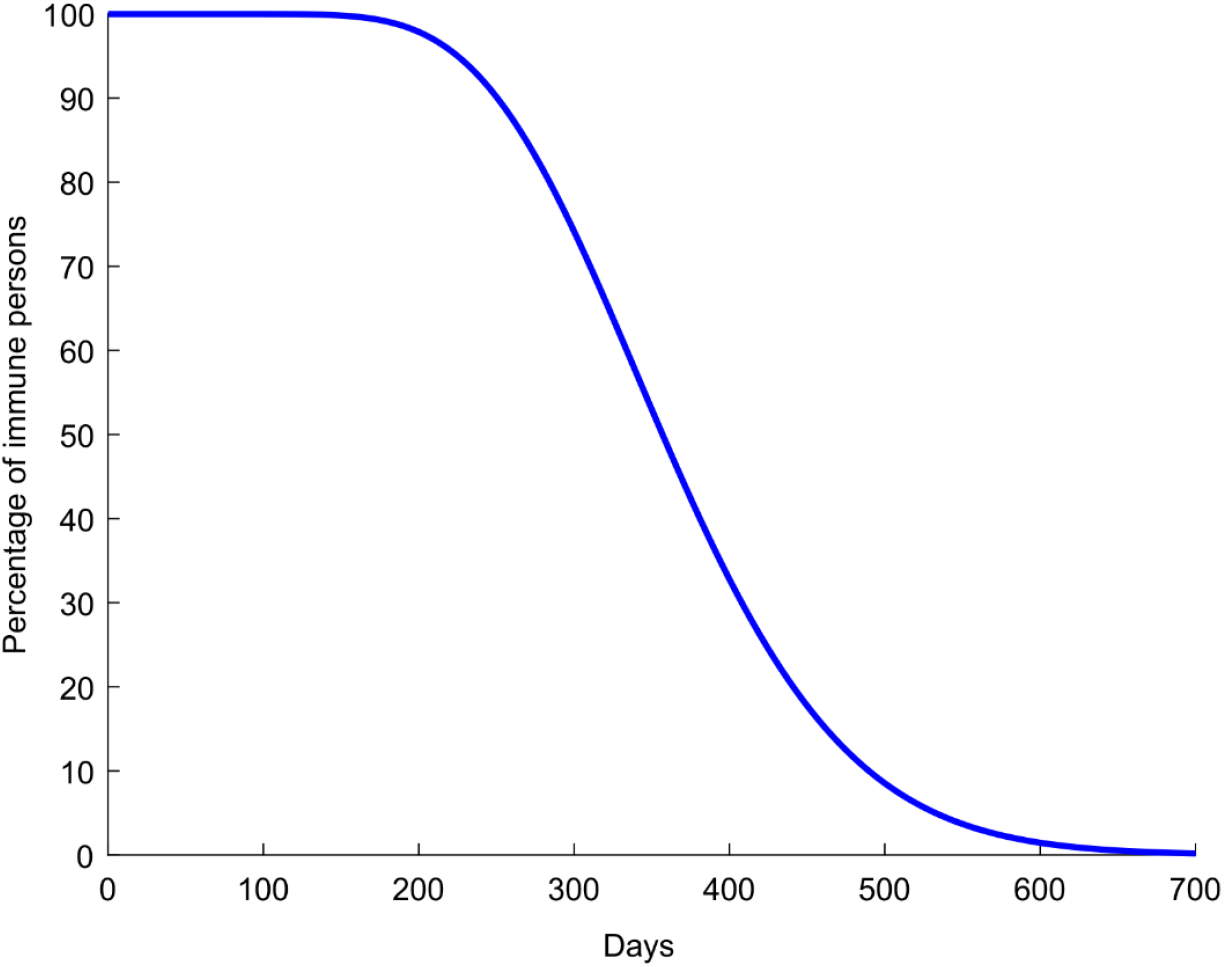
Time course of waning natural infection and vaccine immunity. Waning of natural infection and vaccine immunity was parametrized using a gamma distribution with order *n* = 15. That is, most people lose their immunity after about one year and only a small minority lose it much sooner or much later than one year.

#### C. Parameter values

The input parameters of the model were chosen based on current empirical data for SARS-CoV-2 natural history and epidemiology. The parameter values are listed in Table S2.

**Table S2.** Model assumptions in terms of parameter values.

| Parameter | Symbol | Value | Justification |
| --- | --- | --- | --- |
| Duration of latent infection | $1/\delta$ | 3.69 days | Based on existing estimate [6] and based on a median incubation period of 5.1 days [7] adjusted by observed viral load among infected persons [8] and reported transmission before onset of symptoms [9] |
| Duration of infectiousness | $1/\eta_M$ ;<br>$1/\eta_{DS}$ ;<br>$1/\eta_{DC}$ | 3.48 days | Based on existing estimate [6] and based on observed time to recovery among persons with mild infection [6, 10] and observed viral load in infected persons [8, 9, 11] |
| Duration of severe disease following onset of severe disease | $1/\eta_S$ | 28 days | Observed duration from onset of severe disease to recovery [10] |
| Duration of hospitalization for critical infection | $1/\eta_C$ | 42 days | Observed duration from onset of critical disease to recovery [10] |
| Life expectancy in US | $1/\mu$ | 79.10 years | United Nations World Population Prospects database [12] |
| Life expectancy in China | $1/\mu$ | 76.47 years | |
| Proportion of infections that will progress to be mild or asymptomatic infections | $f_M(a)$ | Determined from<br>$f_M(a) + f_S(a) + f_C(a) = 1$ | Observed proportion of infections that eventually develop mild or asymptomatic in France [10, 13, 14] |
| Proportion of infections that will progress to be infections that require hospitalization in acute care beds | $f_S(a)$ | | The distribution and age dependence of asymptomatic/mild, severe, or critical infections was based on the modeled SARS-CoV-2 epidemic in France [15] |
| Age 0-19 years | $RRS1 \times f_S$ | $RRS1 = 0.1$ | Model-estimated relative risk of severe infection based on the SARS-CoV-2 epidemic in France [15] |
| Age 20-29 years | $RRS2 \times f_S$ | $RRS2 = 0.5$ | Model-estimated relative risk of severe infection based on the SARS-CoV-2 epidemic in France [15] |
| Age 30-39 years | $f_S$ | Reference category<br>($f_S = 0.01$) | Model-estimated based on fitting the SARS-CoV-2 epidemic in France [15] |
| Age 40-49 years | $RRS3 \times f_S$ | $RRS3 = 1.2$ | Model-estimated relative risk of severe infection based on the SARS-CoV-2 epidemic in France [15] |
| Age 50-59 years | $RRS4 \times f_S$ | $RRS4 = 2.3$ | Model-estimated relative risk of severe infection based on the SARS-CoV-2 epidemic in France [15] |
| Age 60-69 years | $RRS5 \times f_s$ | $RRS5 = 4.5$ | Model-estimated relative risk of severe infection based on the SARS-CoV-2 epidemic in France [15] |
| Age 70-79 years | $RRS6 \times f_s$ | $RRS6 = 7.8$ | Model-estimated relative risk of severe infection based on the SARS-CoV-2 epidemic in France [15] |
| Age 80+ years | $RRS7 \times f_s$ | $RRS7 = 27.6$ | Model-estimated relative risk of severe infection based on the SARS-CoV-2 epidemic in France [15] |
| Proportion of infections that will progress to be critical infections | $f_c(a)$ | | The distribution and age dependence of asymptomatic/mild, severe, or critical infections was based on the modeled SARS-CoV-2 epidemic in France [15] |
| Age 0-19 years | $RRC1 \times f_c$ | $RRC1 = 0.21$ | Model-estimated relative risk of critical infection based on the SARS-CoV-2 epidemic in France [15] |
| Age 20-29 years | $RRC2 \times f_c$ | $RRC2 = 0.33$ | Model-estimated relative risk of critical infection based on the SARS-CoV-2 epidemic in France [15] |
| Age 30-39 years | $f_c$ | Reference category<br>$f_c = 0.0002$ | Model-estimated based on fitting the SARS-CoV-2 epidemic in France [15] |
| Age 40-49 years | $RRC3 \times f_c$ | $RRC3 = 1.83$ | Model-estimated relative risk of critical infection based on the SARS-CoV-2 epidemic in France [15] |
| Age 50-59 years | $RRC4 \times f_c$ | $RRC4 = 4.67$ | Model-estimated relative risk of critical infection based on the SARS-CoV-2 epidemic in France [15] |
| Age 50-59 years | $RRC4 \times f_c$ | $RRC4 = 4.67$ | Model-estimated relative risk of critical infection based on the SARS-CoV-2 epidemic in France [15] |
| Age 60-69 years | $RRC5 \times f_c$ | $RRC5 = 10.58$ | Model-estimated relative risk of critical infection based on the SARS-CoV-2 epidemic in France [15] |
| Age 70-79 years | $RRC6 \times f_c$ | $RRC6 = 13.61$ | Model-estimated relative risk of critical infection based on the SARS-CoV-2 epidemic in France [15] |
| | $RRC7 \times f_c$ | $RRC7 = 8.67$ | Model-estimated relative risk of critical infection based on the SARS-CoV-2 epidemic in France [15] |
| Disease mortality rate in each age group | $\alpha(a)$ | | The distribution and age dependence of COVID-19 mortality was based on the modeled SARS-CoV-2 epidemic in France [15] |
| Age 0-19 years | $RRD1 \times \alpha$ | $RRD1 = 0.10$ | Model-estimated relative risk of death based on the SARS-CoV-2 epidemic in France [15] |
| Age 20-29 years | $RRD2 \times \alpha$ | $RRD2 = 0.40$ | Model-estimated relative risk of death based on the SARS-CoV-2 epidemic in France [15] |
| Age 30-39 years | $\alpha$ | Reference category<br>($\alpha = 0.0006$) | Model-estimated based on fitting the SARS-CoV-2 epidemic in France [15] |
| Age 40-49 years | $RRD3 \times \alpha$ | $RRD3 = 3.00$ | Model-estimated relative risk of death based on the SARS-CoV-2 epidemic in France [15] |
| Age 50-59 years | $RRD4 \times \alpha$ | $RRD4 = 10.00$ | Model-estimated relative risk of death based on the SARS-CoV-2 epidemic in France [15] |
| Age 60-69 years | $RRD5 \times \alpha$ | $RRD5 = 45.00$ | Model-estimated relative risk of death based on the SARS-CoV-2 epidemic in France [15] |
| Age 70-79 years | $RRD6 \times \alpha$ | $RRD6 = 120.00$ | Model-estimated relative risk of death based on the SARS-CoV-2 epidemic in France [15] |
| | $RRD7 \times \alpha$ | $RRD7 = 505.00$ | Model-estimated relative risk of death based on the SARS-CoV-2 epidemic in France [15] |
| Overall infectious contact rate | $\beta$ | 0.33 contacts per day | Chosen to yield the desired value of $R_0 = 1.2$ in the US |
| | | 0.28 contacts per day | Chosen to yield the desired value of $R_0 = 1.0$ in China |

**Figure S4.**
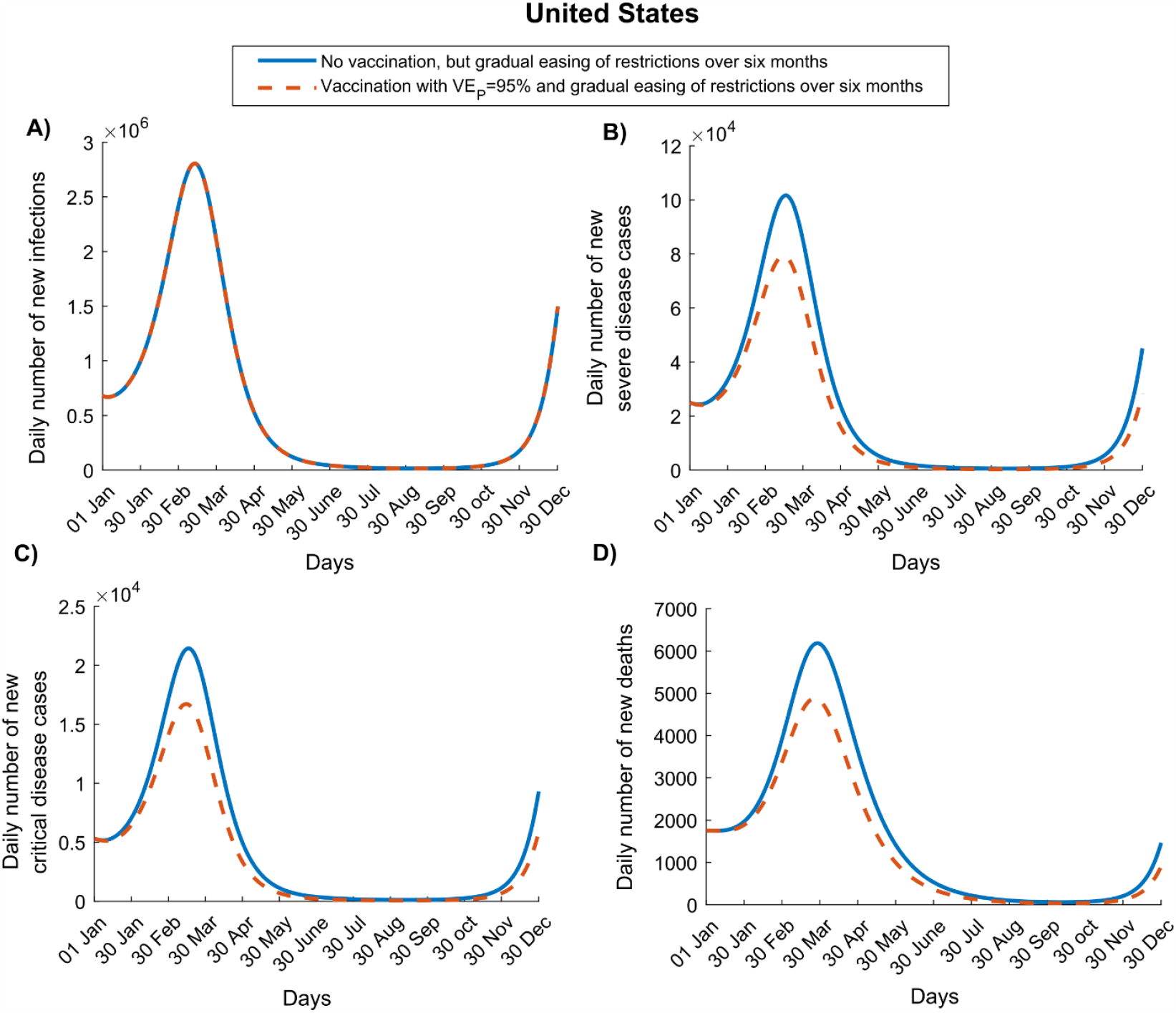
Impact of SARS-CoV-2 vaccination on numbers of A) new infections, B) new severe disease cases, C) new critical disease cases, and D) new deaths in the United States. The vaccine is assumed to have no efficacy against infection, but an efficacy of 95% against severe and critical disease. It is introduced on January 1, 2021, when the cumulative proportion of the population infected is 20%. Vaccine coverage is scaled up to reach 80% by December 31, 2021. The duration of both vaccine protection and natural immunity is one year. This scenario assumes an *R*_0_ of 1.2 on January 1, 2021, which increases with gradual easing of restrictions to reach 4.0 after six months.

**Figure S5.**
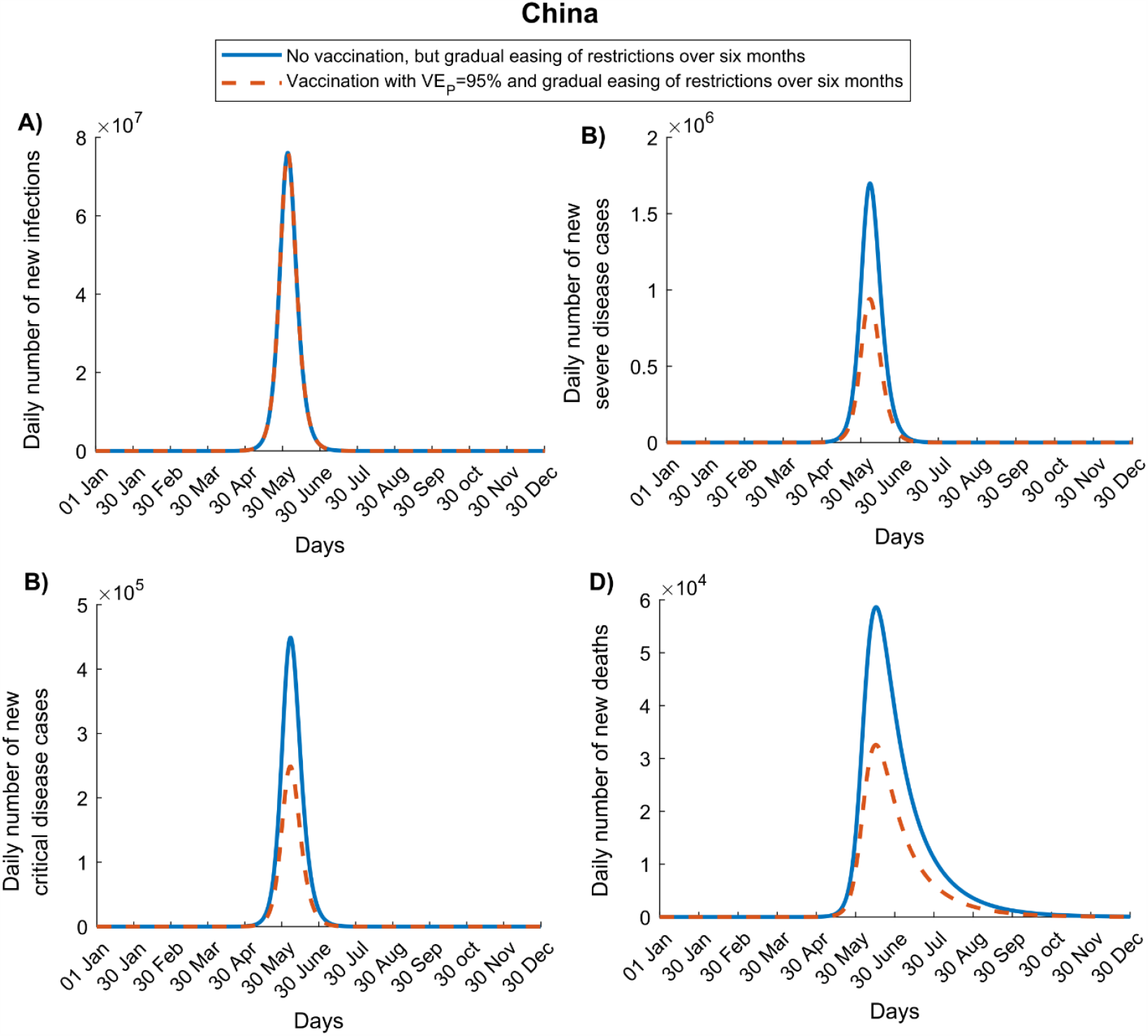
Impact of SARS-CoV-2 vaccination on numbers of A) new infections, B) new severe disease cases, C) new critical disease cases, and D) new deaths in China. The vaccine is assumed to have no efficacy against infection, but an efficacy of 95% against severe and critical disease and is introduced on January 1, 2021. Vaccine coverage is scaled up to reach 80% by December 31, 2021. The duration of both vaccine protection and natural immunity is one year. This scenario assumes an *R*_0_ of 1.0 on January 1, 2021, which increases with the gradual easing of restrictions to reach 4.0 after six months.

**Figure S6.**
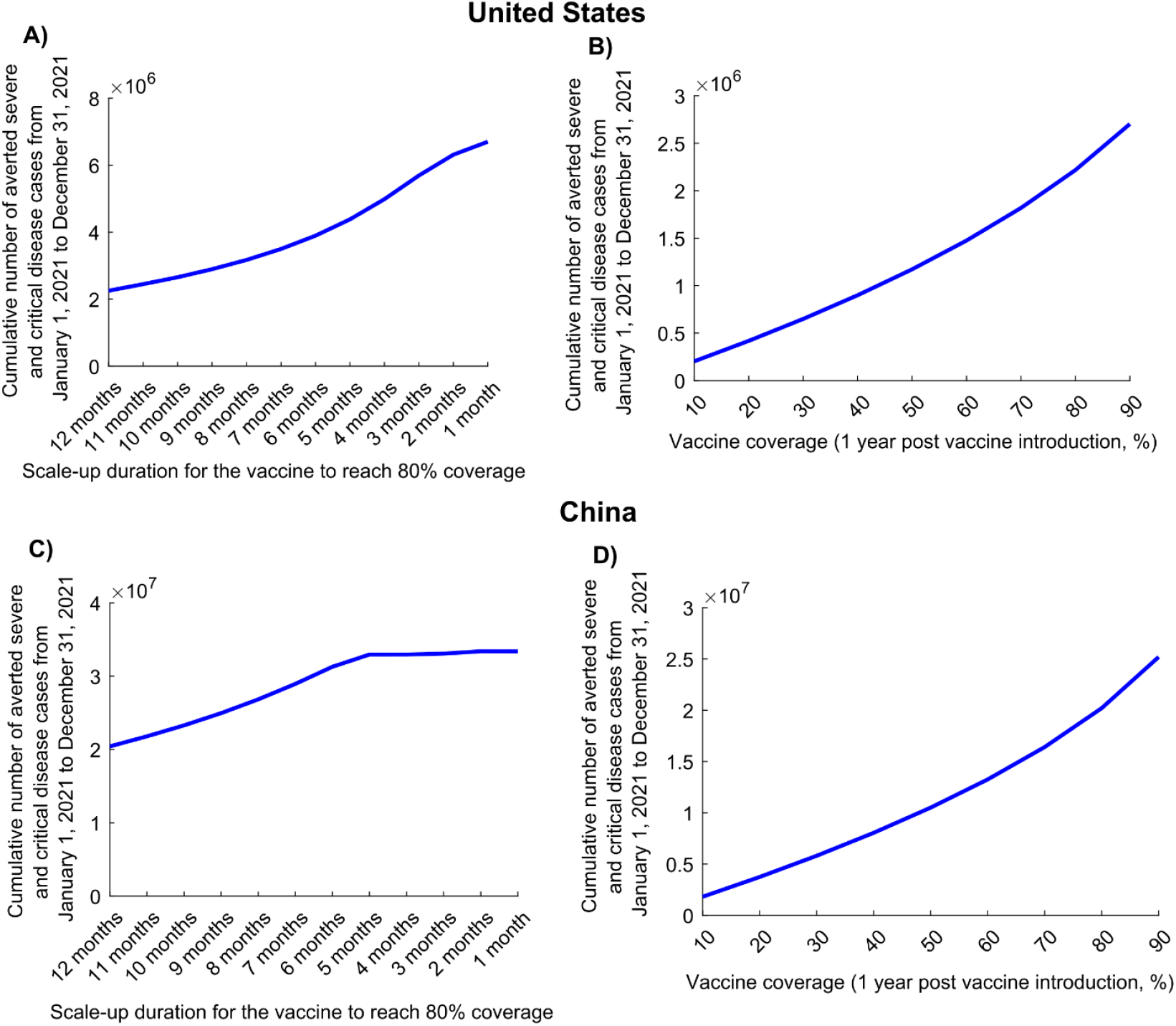
Impact of vaccine scale-up duration and vaccine coverage on numbers of averted severe and critical disease cases for a vaccine that protects only against disease. Cumulative numbers of averted severe and critical disease cases in A) the United States and C) China at different vaccination scale-up intervals to reach 80% coverage. Cumulative numbers of averted severe and critical disease cases in B) the United States and D) China at varying levels of vaccine coverage. The vaccine is assumed to have no efficacy against infection, but an efficacy of 95% against severe and critical disease and is introduced on January 1, 2021, when the cumulative proportion of the population infected is 20% in the United States and 0% in China. The duration of both vaccine protection and natural immunity is one year. This scenario assumes a gradual easing of restrictions within 6 months.

**Figure S7.**
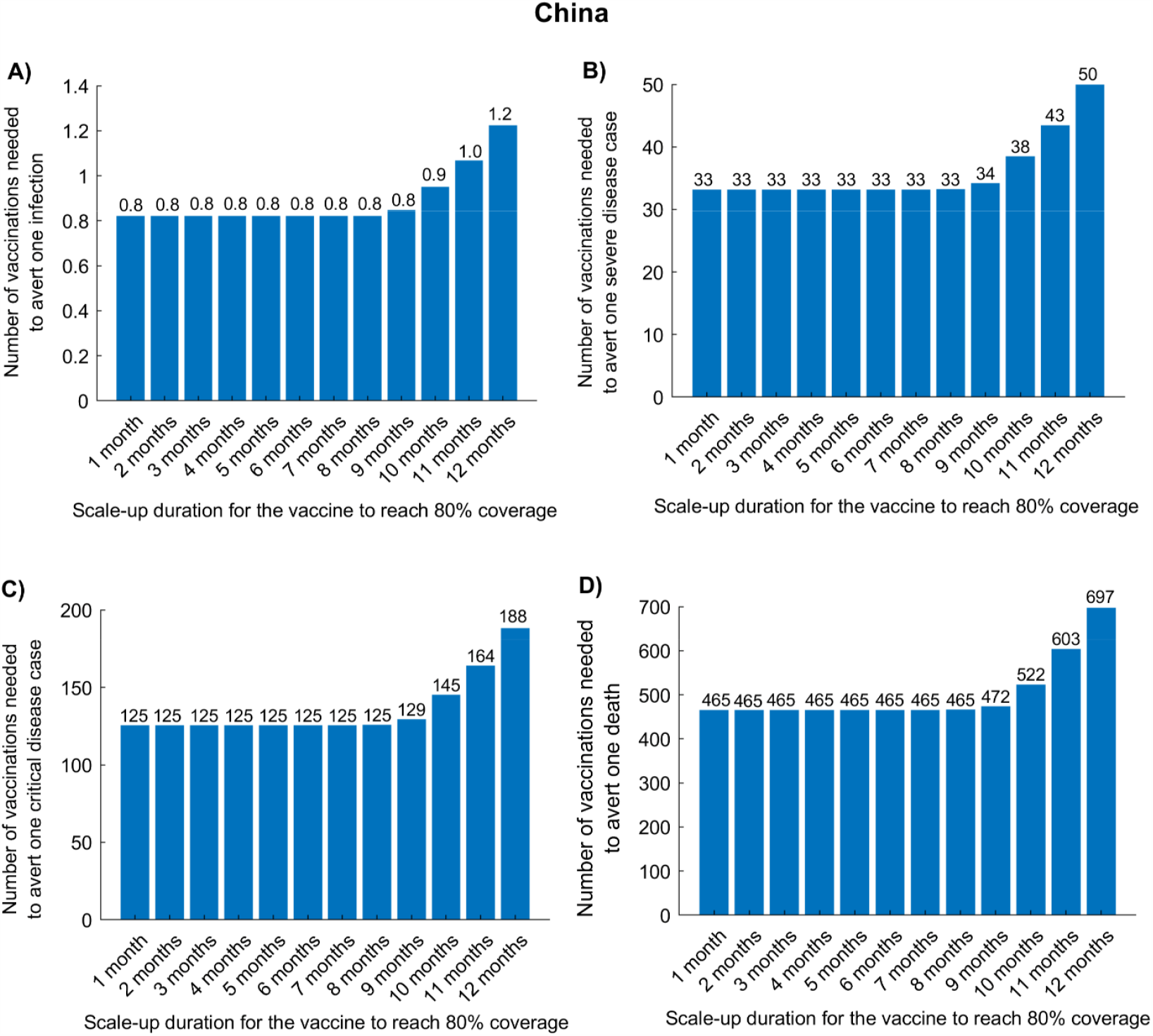
Impact of vaccine scale-up duration on the number of vaccinations needed to avert one infection (A), one severe disease case (B), one critical disease case (C), and one death (D) in China. The vaccine is assumed to have an efficacy of 95% against infection and is introduced on January 1, 2021, when the cumulative proportion of the population infected is 0%. The duration of both vaccine protection and natural immunity is one year. This scenario assumes a gradual easing of restrictions within 6 months.

**Figure S8.**
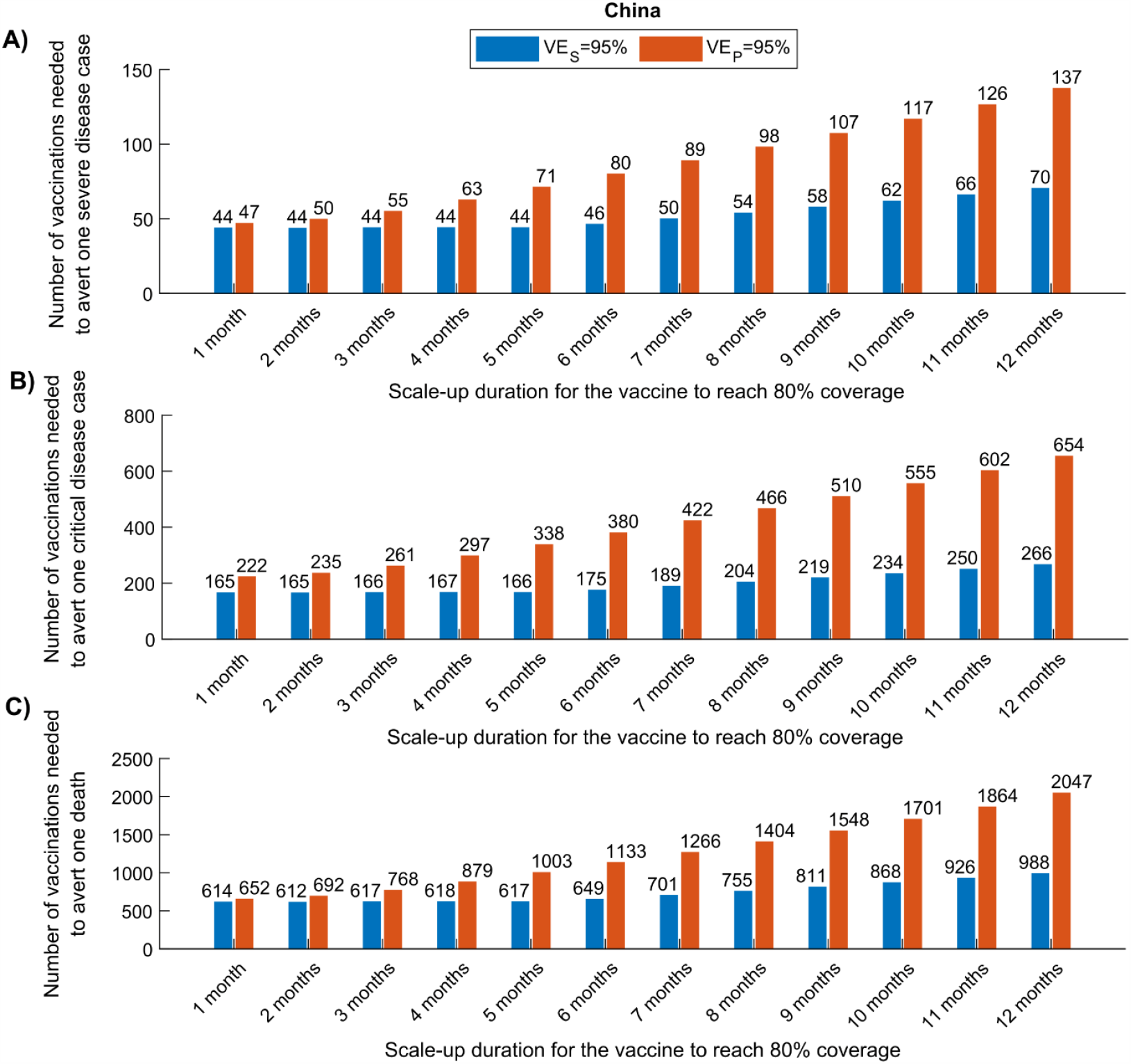
Comparison of the impact of a vaccine acting against infection (*VE*_*S*_ efficacy) versus a vaccine acting only against disease (*VE*_*P*_ efficacy) in China. The number of vaccinations needed to avert one severe disease case (A), one critical disease case (B), and one death (C), for a vaccine with *VE*_*S*_ = 95% versus a vaccine with *VE*_*P*_ = 95%. The vaccine is introduced on January 1, 2021, when the cumulative proportion of the population infected is 0%. The duration of both vaccine protection and natural immunity is one year. This scenario assumes a gradual easing of restrictions within 6 months.

**Figure S9.**
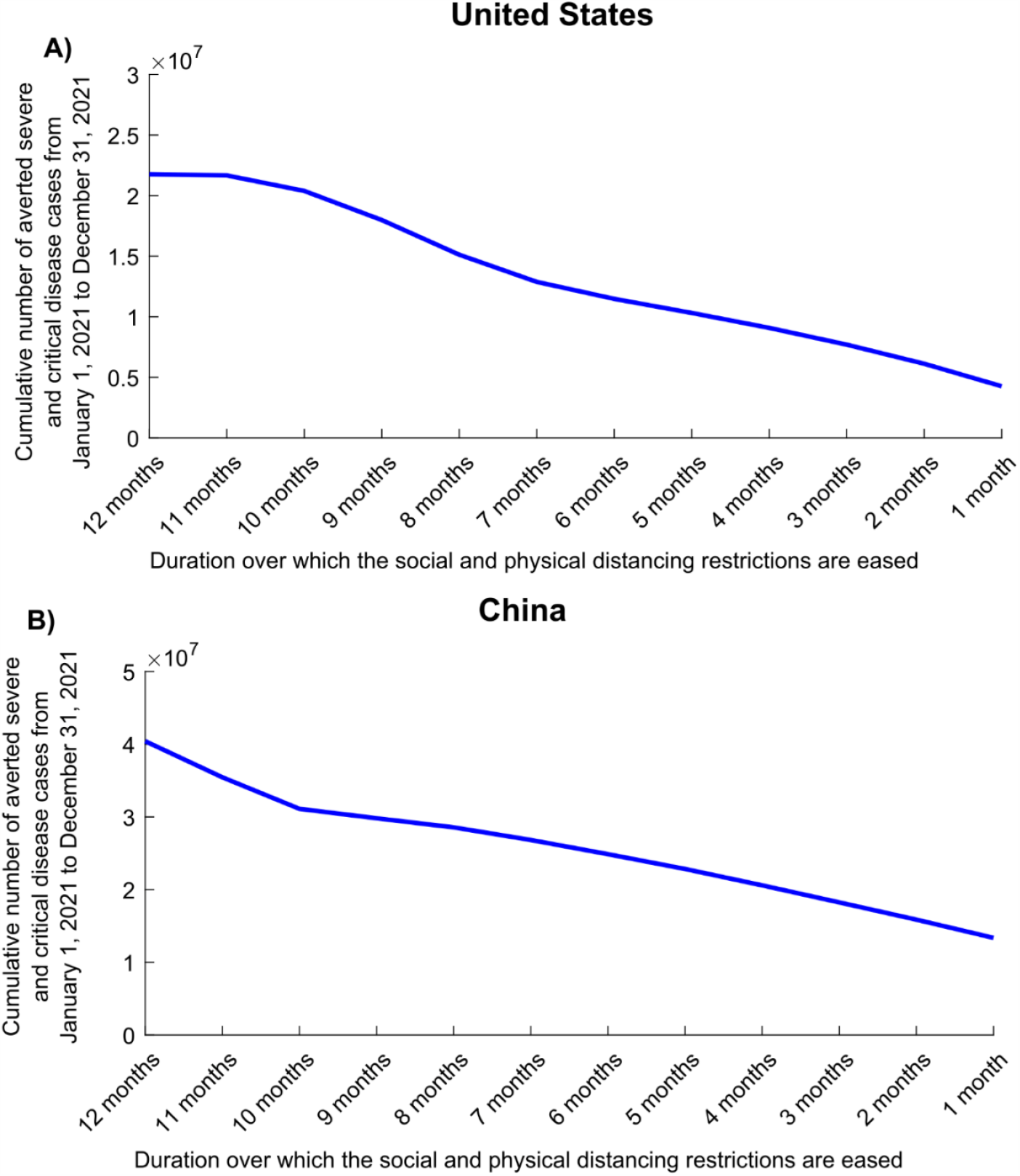
Impact of the duration of easing social and physical distancing restrictions on the number of averted severe and critical disease cases. The cumulative number of averted severe and critical disease cases in A) the United States and B) China at different durations of easing of restrictions. The vaccine is assumed to have an efficacy of 95% against infection and is introduced on January 1, 2021, when the cumulative proportion of the population infected is 20% in the United States and 0% in China. The duration of both vaccine protection and natural immunity is one year.

**Figure S10.**
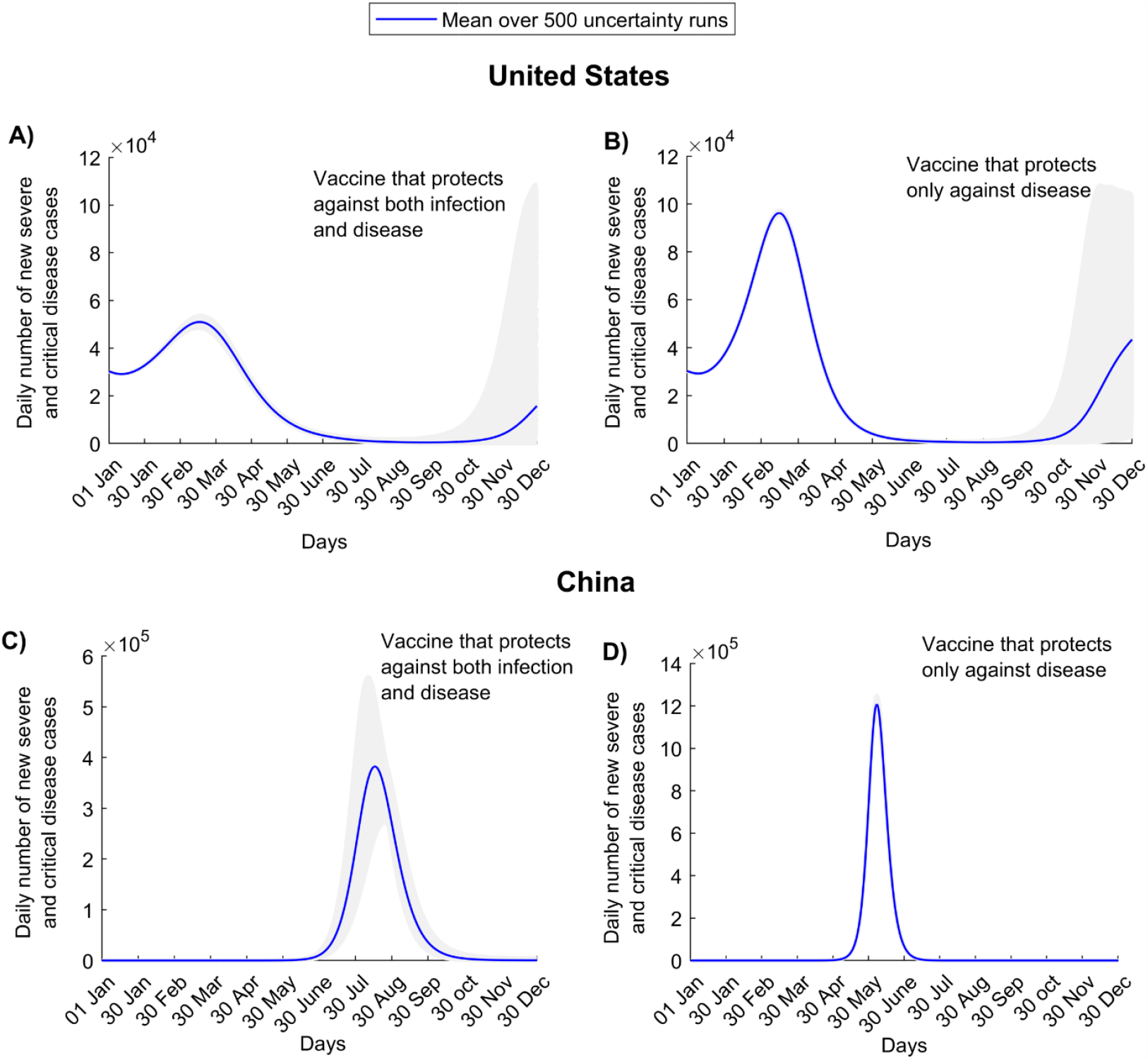
Uncertainty analyses. Numbers of new severe and critical disease cases in the United States assuming A) a vaccine that protects against both infection and disease and B) a vaccine that protects only against disease. Numbers of new severe and critical disease cases in China assuming C) a vaccine that protects against both infection and disease and D) a vaccine that protects only against disease. These scenarios assume gradual easing of restrictions within 6 months. Shaded areas are the results of the 500 uncertainty runs, while the solid line is the mean of those runs.

